# Performance assessment of a new indirect rapid diagnostic test for plague detection in humans and other mammalian hosts

**DOI:** 10.1101/2021.10.19.21265227

**Authors:** Matheus Filgueira Bezerra, Wagner José Tenório dos Santos, Igor Vasconcelos Rocha, Natalia Rocha Nadaes, Filipe Dantas-Torres, Kamila Gaudêncio da Silva Sales, Osvaldo Pompilio de Melo Neto, Marise Sobreira Bezerra da Silva, Edimilson Domingos Silva, Alzira Maria Paiva de Almeida, Christian Robson de Souza Reis

## Abstract

Plague is a flea-borne zoonosis that affects a wide range of mammals and still causes outbreaks in human populations yearly across several countries. While crucial for proper treatment, early diagnosis is still a major challenge in low- and middle-income countries due to poor access to laboratory infrastructure in rural areas. To tackle this issue, we developed and evaluated a new F1-based rapid diagnostic test (RDT) as an alternative method for plague diagnosis in humans and other mammals in the field. In this study, 187 serum samples from humans, dogs, rodents and rabbits were retrospectively assessed using the Plague RDT method. To calculate its performance rates, results were confronted to those obtained by hemagglutination (HA) and ELISA, considered as the reference standards. Remarkably, the results from RDT were in full agreement with those from the ELISA and HA assays, resulting in 100% (CI 95% = 95.5-100%) of sensitivity and 100% (CI 95% = 96.6-100%) of specificity. Accordingly, the Cohen’s Kappa test coefficient was 1.00 (almost perfect agreement). Moreover, the RDT showed no cross-reaction when tested with sera from individuals positive to other pathogens, such as *Yersinia pseudotuberculosis, Yersinia enterocolitica, Anaplasma platys, Erliquia canis and Leishmania infantum*. Although preliminary, this study brings consistent proof-of-concept results with high performance rates of the Plague RDT when compared to other methods well-established in the plague routine serodiagnosis. Although further human and animal population-based studies will be necessary to validate these findings, the data presented here show that the Plague RDT is highly sensitive and specific, polyvalent to several mammal species and simple to use in field surveillance or point-of-care situations with instant results.

## 1 INTRODUCTION

Plague is a flea-borne zoonosis caused by the gram-negative bacterium *Yersinia pestis*, which can infect a wide range of mammals and has been found in human remains that can be traced back to prehistoric ages (Barbieri, 2021; Bramanti, 2016). Although much of its concept as an ancient disease capable of devastating entire societies comes from past pandemics, plague outbreaks still occur yearly in several countries spread across Africa, Asia and the Americas (Valles, 2020). As new plague outbreaks are still possible and *Y. pestis* can be used as a biological weapon, healthcare and surveillance systems must remain prepared for emergencies (Bertherat, 2007).

Plague can be effectively treated with a wide range of antibiotics and to date multidrug resistance is still a rare event in *Y. pestis* (Urich, 2012; Dai, 2021). Due to its aggressive clinical evolution, early detection of the disease is crucial and is considered one of the main challenges faced by health authorities, given the typically remote and rural landscape of plague incidence (Valles, 2020; Demeure, 2019). Moreover, during periods of quiescence in plague foci, serological surveillance of wild and domestic animals must be continuous, as it may identify areas at increased risk for human transmission (WHO, 2006).

Considered as a neglected disease, many endemic low- and middle-income countries have poor access to large-scale plague screening tools. Conventional bacteriological, molecular and serological methods require specialized personnel and laboratory infrastructure and are difficult to implement outside of urban centers (Demeure, 2019). Recently, our group developed a single ELISA test based on a *Staphylococcus aureus* protein A conjugate that is efficient for the serological detection of anti-plague antibodies from a wide range of mammalian species (Bezerra, 2021). This approach, however, still requires a laboratory-based infrastructure. To overcome this barrier and allow point-of-care diagnostics, we now took advantage of the same protein A system to develop a new rapid diagnostic test (RDT) as an alternative method for plague diagnosis in humans and other mammals in the field.

## 2 METHODS

### 2.1 F1 production

The F1 capsular antigen, specific for *Y. pestis*, was extracted from the A1122 *Y. pestis* strain, according to a previously described protocol (Chu, 2000), in a biosafety level 3 facility. The purified F1 antigen was evaluated by western blotting and quantified using the NanoDrop One^C^ Microvolume UV-Vis Spectrophotometer (Thermo-Fischer, USA).

### 2.2 Serum samples

A total of 187 sera well-characterized for anti-plague antibodies (with concordant results in hemagglutination and ELISA) from 46 control rabbits, 43 humans, 44 rodents and 54 dogs were retrospectively accessed in this study. The sera originated from the routine surveillance of domestic and wild animals in the Brazilian plague areas, or from investigations of human plague outbreaks, and are maintained in the plague serum bank of the Plague National Reference Service (SRP) of the Aggeu Magalhães Institute.

To evaluate cross-reactions with other endemic diseases, we included canine sera positive for ehrlichiosis and anaplasmosis (both diagnosed with the SNAP 4Dx Idexx test) as well as leishmaniasis (diagnosed with the DPP® Leishmaniose Canina and ELISA EIE Leishmaniose Visceral Canina Bio-Manguinhos test). Rabbits immunized with *Yersinia pseudotuberculosis* and *Yersinia enterocolitica*, pathogenic to humans, were also evaluated.

### 2.3 Rabbit immunization

The 25 positive control sera were derived from rabbits immunized with formol-killed *Y. pestis* or with the purified F1 antigen, as previously described (Tavares et al., 2020). Three of those were from rabbits immunized with the reference EV76 or A1122 *Y. pestis* strains in independent experiments, 18 were immunized with diverse Brazilian *Y. pestis* strains from the Fiocruz-CYP bacterial culture collection maintained at the Aggeu Magalhães Institute (http://cyp.fiocruz.br), and four were immunized with the F1 antigen. The negative control rabbit sera derived from animals not submitted to any immunization.

### 2.4 Ethics aspects

The immunization of rabbits was approved by the Animal Ethics Committee of the Aggeu Magalhães Institute (CEUA/Fiocruz, protocol number: L-020/09). The use of human serum samples in this research was approved by Ethics Committee of the same institution (CEP/IAM/FIOCRUZ-PE, protocol number: CAAE 50163615.8.0000.5190).

### 2.5 Rapid diagnostic test

For the immunochromatographic test, the F1 antigen and the reaction control (protein A) were dispensed side by side onto nitrocellulose membranes. The impregnated membrane was then assembled in an adhesive card as were the sample, the conjugate and the waste pads (Silva, 2020). The adhesive card was then cut into strips, which were individually placed inside a plastic cassette designed for lateral flow tests. Once assembled, the test is performed by adding serum (5 µL), followed by the addition of three drops of running buffer. The buffer allows the sample to flow across the length of the nitrocellulose membrane strip. Results were read visually after 15 minutes of incubation at room temperature (mean, 22°C). Positive results are characterized by the appearance of colored bands (pink/purple) in the test (F1) and control lines, whereas negative results showed only the control line. If the control line did not appear, the test was considered invalid.

### 2.6 Hemagglutination and ELISA

The hemagglutination (HA) assay was performed according to Chu et al. (2000). In brief, F1-coated sheep red blood cells were incubated with the test serum serially diluted in eight wells in HA buffer (0.85% saline + normal rabbit serum), starting from the 1/4 dilution. The specificity of HA was accessed by the hemagglutination inhibition assay (HI), adding 100 µg/mL of the soluble F1 antigen to the HA buffer. The test was considered positive when the HA endpoint is depressed by three or more HI dilutions (titers ≥ 1/16 are considered positive).

The ELISA-protein A assay followed the protocol previously described by Bezerra et al. (2021). In short, 96-well plates were incubated overnight with 250 ng per well of F1 diluted in 0.05 M, pH 9.6 carbonate-bicarbonate buffer. Next, the plates were washed and blocked with 10% low fat milk/PBS solution. Samples were incubated in a dilution of 1:500 in milk/PBS-Tween (1%) and washed again. Next, a 1:10.000 protein A–peroxidase conjugate solution was incubated at room temperature and washed with PBS-Tween, developed with ortho-phenylenediamine dihydrochloride (OPD)/H_2_O_2_ and interrupted with sulfuric acid (H_2_SO_4_). Plates were read at the optical density of 490 nm (iMark™ Microplate Absorbance Reader, Bio-Rad, USA). All samples were measured in triplicate and the cut-off point was determined as described elsewhere (Bezerra et al., 2021).

### 2.7 Statistical analysis

Serum samples previously characterized and with concordant results in HA and ELISA testing were used as the reference standards to calculate the Plague RDT performance rates. Sensitivity, specificity and confidence intervals were calculated using the MedCalc statistical software (https://www.medcalc.org). The Kappa test was calculated using the QuickCalcs GraphPad tool (https://www.graphpad.com/quickcalcs/kappa2) and interpreted as following: poor agreement (k = 0), slight agreement (0.20 ≤ k > 0), fair agreement (0.40 ≤ k ≥ 0.21), moderate agreement (0.6 ≤ k ≥ 0:41), substantial agreement (0.80 ≤ k ≥ 0.61), and almost perfect agreement (1.0 ≤ k ≥ 0.81). Statistical tests were applied with a 95% confidence interval.

## 3 RESULTS

To assess the performance of the new F1-based Plague RDT, we initially analyzed control sera obtained from rabbits inoculated with formol-killed *Y. pestis* or F1 antigen (positive controls, n=25) or uninoculated rabbits (negative controls, n=21). Visual analysis of RDTs showed the presence of both test and control bands for all positive control samples and only the control band for all negative control samples (**Figure 1A**), matching their inoculation status. Next, we expanded our analysis to well-characterized sera from humans (n=43), rodents (n=44) and dogs (n=54), which were assigned as positive or negative according to results from ELISA-protein A and hemagglutination (HA) assays (**Figure 1B**), standard methods used in the routine plague diagnosis (total = 187 samples). Remarkably, the results from RDT were in full agreement with those from the ELISA and HA assays (**Figure 1C**), with 100% (CI 95% = 95.5-100%) of sensitivity and 100% (CI 95% = 96.6-100%) of specificity. Accordingly, the Cohen’s Kappa test coefficient was 1.00 (i.e., almost perfect agreement). These findings were reproduced in the analyses for each individual species, as shown in Table 1.

**Table 1.**
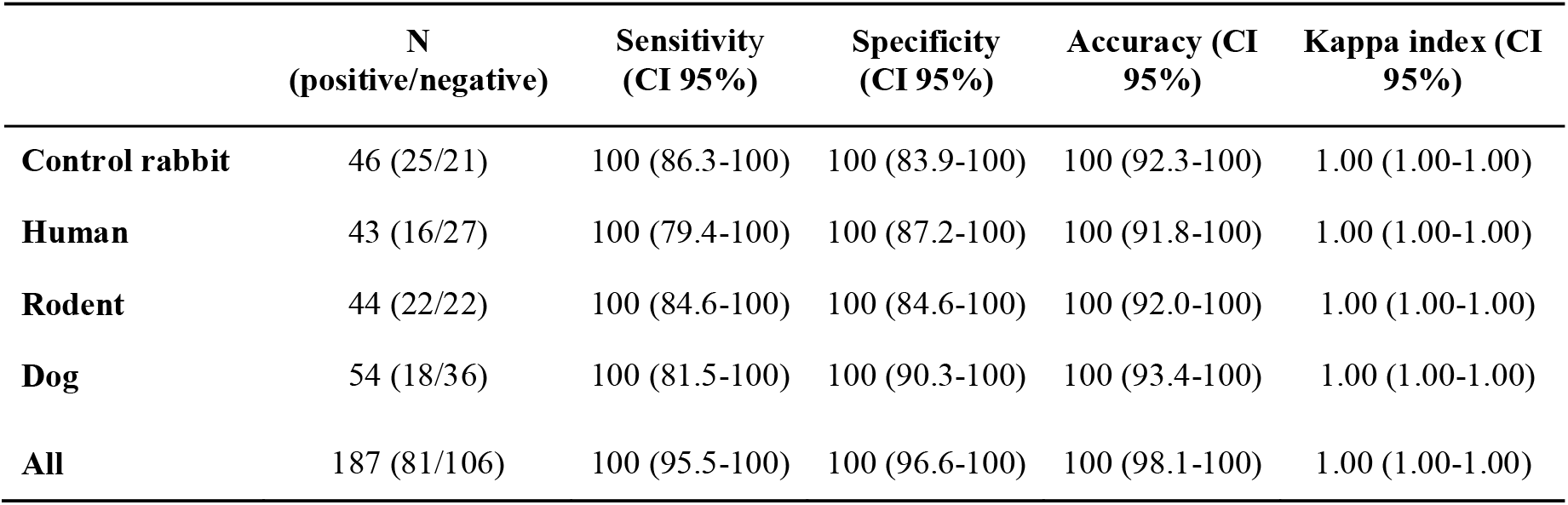
Performance assessment of the Plague RDT according to results from the ELISA and HA reference methods.

|  | N<br>(positive/negative) | Sensitivity<br>(CI 95%) | Specificity<br>(CI 95%) | Accuracy (CI<br>95%) | Kappa index (CI<br>95%) |
| --- | --- | --- | --- | --- | --- |
| **Control rabbit** | 46 (25/21) | 100 (86.3-100) | 100 (83.9-100) | 100 (92.3-100) | 1.00 (1.00-1.00) |
| **Human** | 43 (16/27) | 100 (79.4-100) | 100 (87.2-100) | 100 (91.8-100) | 1.00 (1.00-1.00) |
| **Rodent** | 44 (22/22) | 100 (84.6-100) | 100 (84.6-100) | 100 (92.0-100) | 1.00 (1.00-1.00) |
| **Dog** | 54 (18/36) | 100 (81.5-100) | 100 (90.3-100) | 100 (93.4-100) | 1.00 (1.00-1.00) |
| **All** | 187 (81/106) | 100 (95.5-100) | 100 (96.6-100) | 100 (98.1-100) | 1.00 (1.00-1.00) |

**Figure 1.**
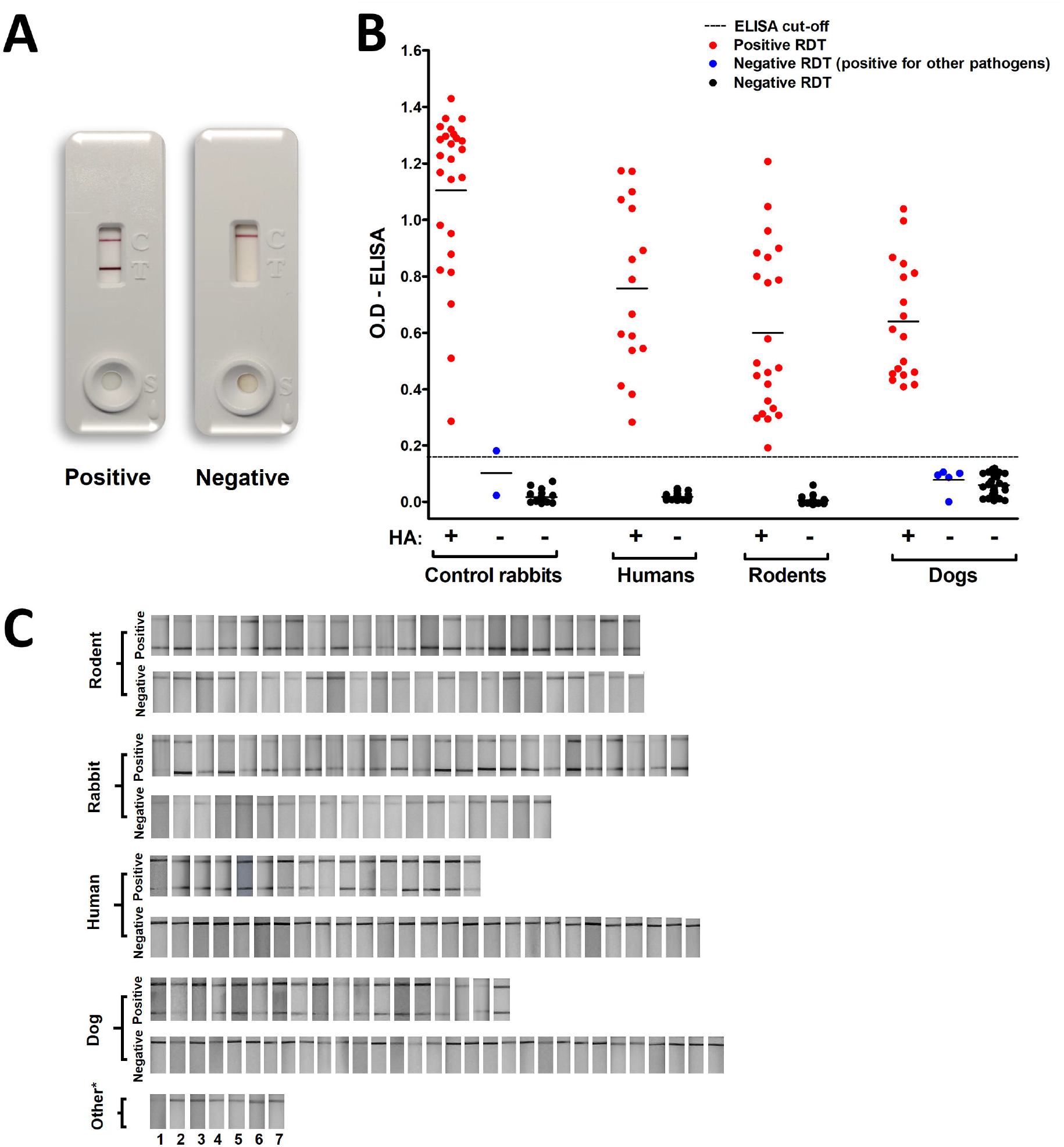
Evaluation of the Plague RDT using positive and negative samples from multiple hosts. Visual profile of a positive and a negative sample (A); Comparison between results from the Plague RDT and the reference methods ELISA and HA (B) and compilation of all tested sera. 1 = Y. enterocolitica, 2 = Y. pseudotuberculosis, 3-4 = anaplasmosis, 5-6 = ehrlichiosis and 7 = leishmaniasis (C).

Due to the central role of serosurveillance of dogs in the monitoring of plague foci, we also evaluated whether sera from dogs infected with other endemic diseases in Brazil, such as anaplasmosis (n=2), ehrlichiosis (n=2) and leishmaniasis (n=1), would cross-react in this test. Additionally, we also tested sera from rabbits immunized with two other Yersiniae pathogenic to humans: *Y. pseudotuberculosis* (n=1) and *Y. enterocolitica* (n=1). No cross-reaction was observed for any of the pathogens tested.

## 4 DISCUSSION

Although preliminary, this study brings consistent proof-of-concept results with high performance rates of the Plague RDT when compared to other methods well-established in the plague routine serodiagnosis. These findings were reproducible in all four distinct groups analyzed herein: humans, rodents, rabbits and dogs. In addition to its high sensitivity and specificity rates, the RDT showed no cross-reaction when tested with sera from dogs positive to other endemic pathogens.

The new Plague RDT tackles two of the main challenges faced in plague diagnosis: the poor access to laboratorial infrastructure at the primary healthcare level in plague foci and the need for tests capable of detecting anti-plague immunoglobulins from a wide range of host species (Demeure, 2019). Beyond its application in the diagnosis of human plague and continuous monitoring of the sentinel animals (Sousa, 2017), this multi-species Plague RDT can be also useful to study the impact of the dispersion of the bacterium across distinct ecosystems (Bevins, 2021). Although direct antigen detection rapid tests are already available in some countries and are very useful for the detection of active disease (Rajerison, 2020; Hsu, 2018), they are not ideal for the surveillance of previously exposed individuals from sentinel animals.

It is important to highlight that some relevant questions still need to be further addressed in a larger study. Due to the current absence of human or animal cases of plague in Brazil, only retrospective samples were assessed herein. Therefore, we were not able to estimate the positive/negative predictive values nor measure the test performance in multiple timepoints considering the onset of symptoms. Nevertheless, taking into consideration that other indirect serologic approaches are reported to have an immunological window for the detection of anti-F1 antibodies of five to eight days (Rasoamanana 1997, Andrianaivoarimanana 2020, Butler 1977), we presume that the Plague RDT most likely would perform within a similar time frame. Moreover, as the Plague RDT takes advantage of the protein-A polyvalence to immunoglobulins from multiple mammalian species, another point to be further evaluated is the performance of the Plague RDT test in other wild species known to be susceptible to plague.

Altogether, the data presented here show that the Plague RDT is highly sensitive and specific, polyvalent to several mammalian species and simple to use in field surveillance or point-of-care situations with instant results.

## Data Availability

All data produced in the present study are available upon reasonable request to the authors

## 5 ACKNOWLEDGMENTS

We are thankful to the Brazilian National Plague Reference Service staff for providing the serum samples to this study and to the Bio-manguinhos staff for providing the equipment to assemble the RDTs.

